# Relationship between early childhood non-parental childcare and diet, physical activity, sedentary behaviour, and sleep: A systematic review of longitudinal studies

**DOI:** 10.1101/19005413

**Authors:** Silvia Costa, Sara E. Benjamin Neelon, Eleanor Winpenny, Veronica Phillips, Jean Adams

## Abstract

**Background:** The rising prevalence of childhood obesity is a global public health concern. Evidence suggests that exposure to non-parental childcare before age six years is associated with increased risk of obesity, diet, and activity behaviours (physical activity, sedentary behaviour, and sleep). However, findings are inconsistent and mostly from cross-sectional studies, making it difficult to identify the direction of causation in associations. This review identified and synthesised the published research on longitudinal associations between non-parental childcare during early childhood, diet, and activity behaviours.

**Methods:** Seven databases were searched using a predefined search strategy. Results were independently double-screened through title/abstract and full-text stages according to predefined criteria. Included studies were tabulated, and evaluated for risk of bias using the Nutrition Evidence Library Bias Assessment Tool.

**Results:** Of 18793 references screened, 13 studies met eligibility criteria and were included in the review. Eight studies reported on diet and seven studies reported on activity behaviour outcomes (three on physical activity, three on sedentary behaviour, and one on sleep). These studies included results on 89 tested childcare:outcome associations. Of 63 associations testing diet outcomes, 37 (59%) were null, and the remainder showed inconsistent patterns. There was an indication of a potential benefit of Head Start providers (vs other care, including parental) on dietary behaviours. Of 26 associations testing activity behaviour outcomes, 22 (85%) were null, and the remainder were inconsistent. Most studies (92%) did not use (or did not report using) valid and reliable outcome measures, and outcome assessors were not blinded (or it was unclear if they were blinded) to children’s exposure status (77%).

**Conclusions:** The scarce available literature indicates little and mixed evidence of a longitudinal association between exposure to non-parental childcare before age six years and diet or activity behaviours. This reflects a paucity of research, rather than clear evidence of no effect. There is an urgent need for studies investigating the longitudinal associations of non-parental childcare on diet and activity behaviours to assess potential lasting effects and mechanisms. Studies should assess whether and how associations vary by provider and child sub-groups, as well as differences by intensity and duration of care.

## BACKGROUND

High rates of childhood obesity are a worldwide concern (1, 2). Globally in 2016, approximately 40.6 million (6%) children under 5 years of age were overweight or obese (1), an increase of over one third from 30.4 million in 2000 (1). Obesity during childhood is associated with increased risk of both obesity and a range of other conditions later in life, including low self-esteem, high blood pressure, insulin resistance, coronary heart disease and stroke (3-5). The early years (<6 years of age) have been repeatedly highlighted as a critical period for the development and prevention of obesity (4, 6, 7), as well as the establishment of related healthy habits, such as healthy diet, physical activity, and sleep patterns (8, 9). Several individual, inter-personal and environmental factors influence the development of childhood obesity (10). Because they affect large numbers of children, environmental factors such as childcare settings represent potential targets for obesity prevention (11, 12). An increasing number of children now attend non-parental childcare prior to 6 years of age, and many spend large proportions of their week days in such care (13, 14). A report by the United Nations’ Children’s Emergency Fund shows that roughly 80% of 3–6 year olds and 25% of 0–3 year olds in developed countries spend time in some form of childcare (14). A growing body of research, including a number of systematic reviews, suggests that attendance at childcare is associated with increased adiposity or risk of obesity in children (15-17). However, the available evidence is inconsistent (15, 16, 18, 19), and may partly depend on different aspects of the childcare received, such as the type (i.e., informal or formal care) or intensity (e.g., number of hours per week).

The pathways through which non-parental childcare might affect obesity are poorly understood (20, 21). Different types and characteristics of childcare settings may have different influences on the development of obesity-related risk factors, such as diet and activity behaviours (including physical activity, sedentary behaviour, and sleep) (12, 22-25). Evidence suggests that some types of non-parental childcare (e.g., grandparents or Head Start in the US) and staff behaviours (e.g., giving non-food rewards and allowing children to self-serve) are associated with diet patterns and behaviours (24, 26). Similarly, different types (e.g., home-based versus centre-based settings) and features (e.g., staff behaviours like playing with children) of childcare are associated with physical activity (27-30) and sedentary behaviour (23, 30, 31) in young children. There is also some evidence that attending some types of childcare is associated with problematic sleeping patterns in young children (22, 25, 32). However, the direction of these associations is mixed, and associations are not consistently found in all population sub-groups or studies. Additionally, the vast majority of the current evidence comes from cross-sectional studies, which makes it difficult to determine the direction of causation. The aim of this review was to systematically gather and synthesise the published research on the longitudinal relationship between non-parental childcare in the early years and diet, physical activity, sedentary behaviour, and sleep. We focused exclusively on longitudinal studies to increase confidence that any association we find might be causal.

## METHODS

This review was part of a larger programme of systematic reviews (including obesity and stress outcomes alongside the diet and activity behaviour outcomes reported here) that was registered with the PROSPERO database (registration number CRD42015027233) (33), and reported in line with the Preferred Reporting Items for Systematic Reviews and Meta-analysis (PRISMA) recommendations (34). The protocol for the overall programme of systematic reviews has been published elsewhere (35).

### Search Strategy

Seven electronic bibliographic databases were searched in January 2016, using a predefined search strategy: MEDLINE, EMBASE, PsycINFO, Web of Science, Scopus, Applied Social Sciences Index and Abstracts (ASSIA), and the Scientific Electronic Library Online (SciELO). Searches were restricted to human subjects, but there were no restrictions placed on publication date or language. The search strategy was based on the key themes of relevance to the overall review as described in the protocol – childcare, adiposity and body mass, physical activity, sedentary behaviour, sleep, diet, and stress – and informed by search strategies of relevant previous systematic reviews (36). An experienced university librarian (VP) reviewed the search strategy, adapted it for different databases, and ran the searches. An example of the search strategy used for the MEDLINE and Embase databases can be found in Supplement 1. Results were merged from the different databases and managed using EndNote® software. The searches were re-run at the end of May 2017.

### Study Selection and eligibility criteria

After removal of duplicates, records were screened in two phases using a pre-piloted procedure (Box 1). In phase one, title and abstracts were screened by two reviewers working independently against the five phase one eligibility criteria described in Figure 1. The full texts of all studies identified by either reviewer as potentially eligible were retrieved. In phase two, full texts were screened by two reviewers working independently against the seven phase two eligibility criteria described in Figure 1. In cases of uncertainty or discrepancy between reviewers, we consulted a third reviewer and consensus was achieved by discussion.

Figure 1 – Eligibility criteria

The number of papers included and excluded at each stage of the review process can be seen in the PRISMA diagram presented in Figure 2. The diagram shows overall number of references screened and excluded (by reason) instead of number of references by outcome of interest, because eligibility at full-text screening (phase two) was done for all six outcomes of interest in the registered protocol for the programme of systematic reviews (obesity, stress, diet, physical activity, sedentary behaviour and sleep) (33). Concurrently, the final number of included studies are presented for behaviour outcomes only that were the focus of the current review – diet, physical activity, sedentary behaviour, and sleep.

Figure 2 – PRISMA diagram.

Details of and justification for each eligibility criteria are described in full in the protocol (35). Studies were included where participants were children aged <6 years and not in primary school at first assessment, and living in middle- and high-income countries as defined by the World Bank (37). Only observational longitudinal study designs, including case-control, prospective, and retrospective designs, were included. The exposure of interest was non-parental childcare where there was between-child variation in exposure, for example by timing of attendance (i.e., age when care started and stopped), intensity (i.e., full-or part-time care), duration (i.e., years of childcare), types (i.e., formal or informal; private or public), or simply attendance versus non-attendance. Studies were included where outcomes were objectively assessed or proxy/self-reported measures of diet, physical activity, sedentary behaviour, or sleep. Studies were excluded if they were not published in peer-reviewed journals.

When the team could not resolve issues of whether eligibility criteria were met, study authors were contacted via email for clarification. If authors did not reply by the end of the data extraction stage, studies were excluded from the review. Conference abstracts, masters and doctoral theses were excluded, as these do not necessarily go through a formal peer-review process. Nevertheless, the authors of any potentially relevant records of these types of references were contacted via email to determine if peer-reviewed journal articles had resulted, and screened articles identified through this route according to the phase two process above.

### Data Extraction and Management

A standardised and pre-piloted form was used to extract data from included studies for assessment of study quality and evidence synthesis. This captured information about study setting and population, exposure and outcome variables, statistical analyses and results, as stated in the protocol (35). The first author extracted these data into an Excel® database, and a second author (JA/SBN) independently checked the extracted information against the full-texts of included studies.

### Data Synthesis

Key information was tabulated (e.g., sample characteristics, exposure, and outcome measures) for each study, grouped by outcome variable, and performed a narrative synthesis of the included studies. Because of heterogeneity in exposure and outcome variables, it was not appropriate to perform a meta-analysis. This also meant that it was not possible, as originally planned, to perform a quantitative synthesis of differences in effect between different types and features of childcare, different outcomes, high- and middle-income countries, ages at exposure, and socio-demographic sub-groups (e.g., by ethnicity). Instead, and because of the sometimes large number of relevant exposure and outcome variables used in included studies, all individual relevant associations reported in the included studies were included here.

### Quality Assessment

An adaptation of the United States Department of Agriculture’s Nutrition Evidence Library Bias Assessment Tool (NEL-BAT) (38) was used to assess risk of bias in included studies. This tool assesses risk of selection, performance, detection, and attrition bias. For observational studies, responses to each of the 13 questions are scored 0–2 (possible range of scores: 0–26), where lower scores indicate lower risk of bias. SC and JA independently assessed all included studies for risk of bias, and disagreements in scores were resolved by discussion.

## RESULTS

The literature search identified 47 529 articles. After de-duplication, 18 793 articles underwent title/abstract screening, and the full texts of 175 articles were reviewed. Thirteen studies (39-51) met all of the eligibility criteria, and were included in the review. Of these, eight studies reported on diet (39, 45-51), three reported on physical activity (39, 41, 44), three on sedentary behaviour (41-43), and one on sleep (40) outcomes. Some studies reported on more than one of diet, physical activity, sedentary behaviour and sleep.

### Summary of Included Studies

A detailed description of each study’s characteristics can be seen in Table 1. Most included studies were from high-income countries, with seven originating from the United States (39, 40, 42, 43), one from Australia (41), one from New Zealand (44), and one from the UK (48). Samples were generally balanced with relation to children’s gender (although three studies did not report gender composition of the sample) (47, 48, 51), but varied greatly both in size (between 34 and 18 050 subjects) and ethnic composition (between 0% and 87% white, with one study not reporting race/ethnicity or country of birth (41), and five providing information only for country of birth) (45, 46, 48, 49, 51).

**Table 1.**
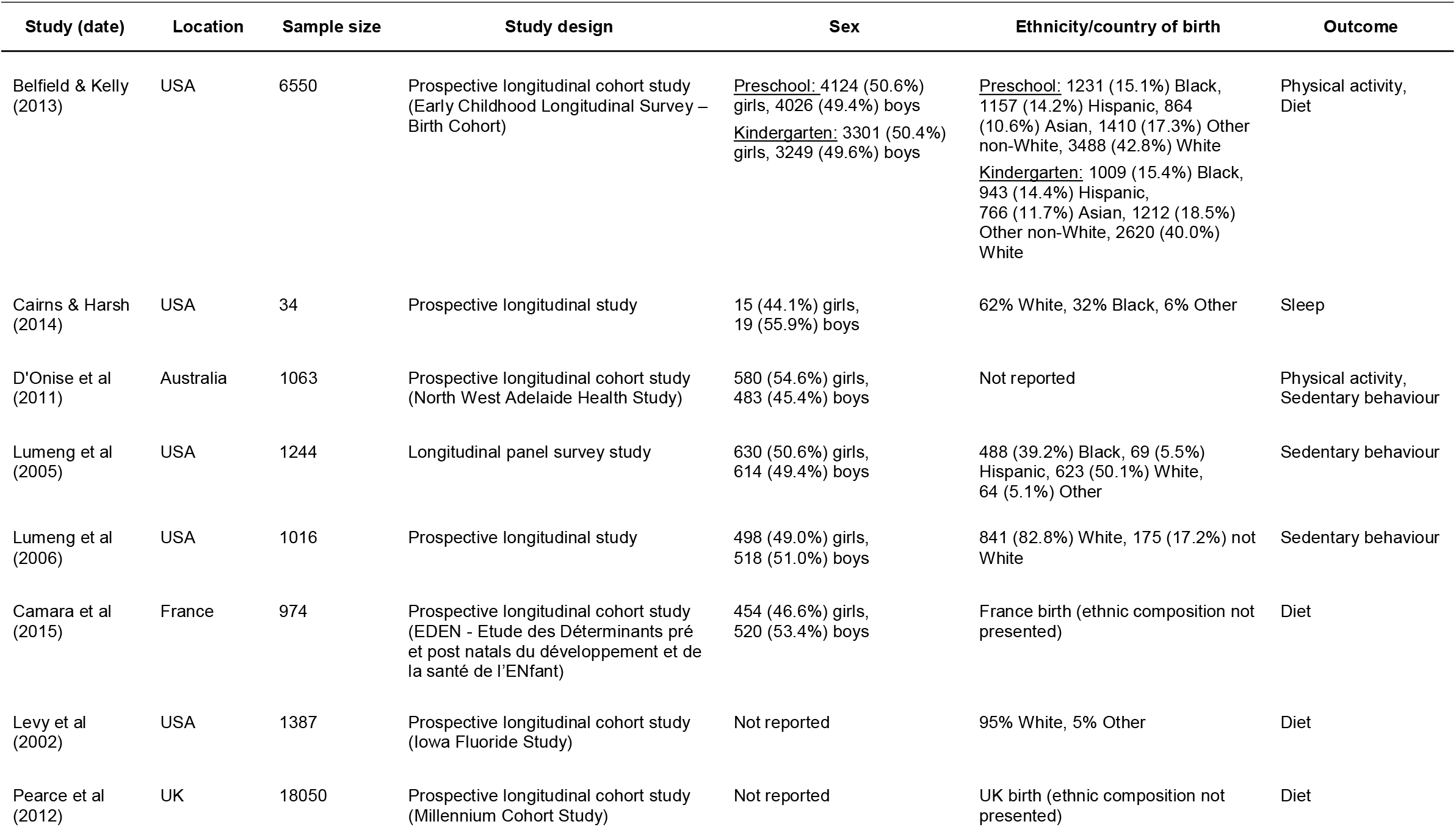
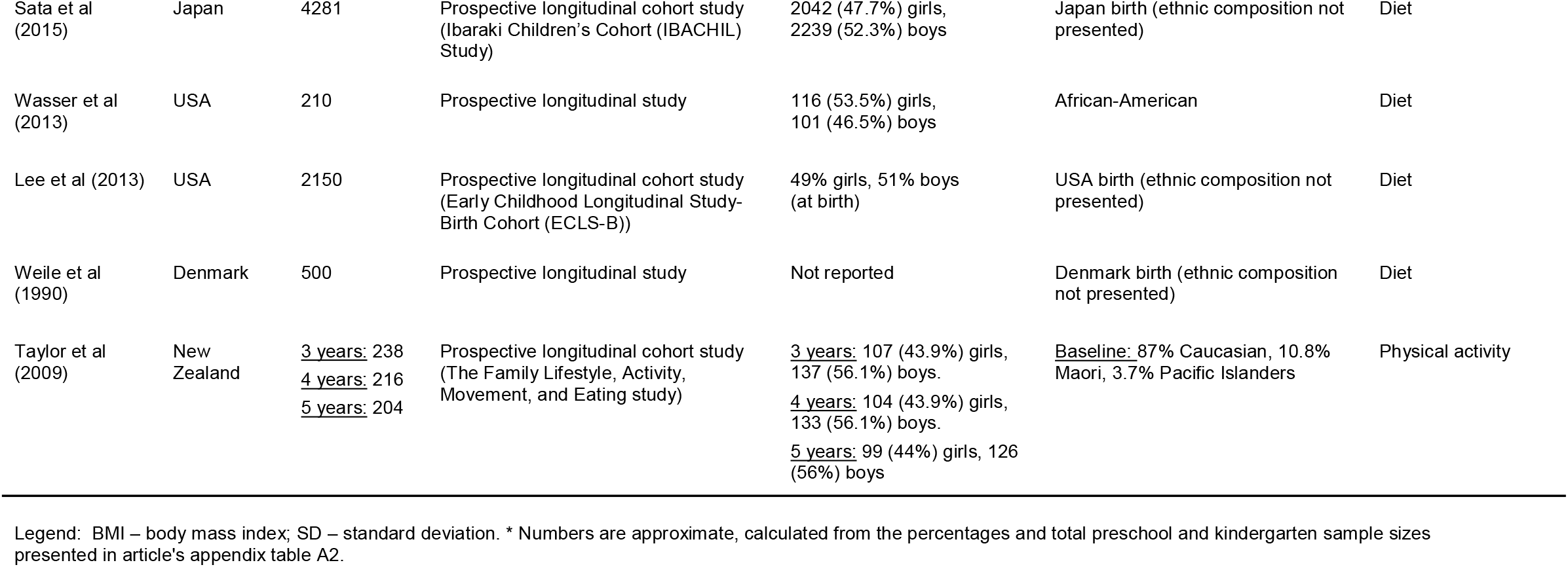
Description of included studies.

All studies assessed exposure to non-parental childcare between birth and 5 years. Measurement of exposure was by proxy-report from a parent or primary caregiver in all studies. Nine studies described childcare exposure in simple categorical terms (e.g., centre-based preschool or Head Start centre versus other/mixed care, including parents) (39-41, 45, 46, 48-51). One study assessed duration of exposure (centre-based preschool for at least 2 years versus other/mixed care, including parents) (39). Five studies assessed intensity of non-parental childcare (e.g., average number of hours in childcare between 24-36 months) (42-44, 47, 48).

Age at outcome assessment varied from 1-12 months to 51 years. For diet outcomes, all studies used proxy-report by a mother or main caregiver (39, 45-51). For physical activity outcomes, most studies used proxy-or self-report (39, 41, 44), with the exception of average accelerometer counts/minute in one case (44). All studies assessing sedentary behaviour outcomes (e.g., hours/day of television (TV) viewing) using proxy-report by a parent. For sleep, quantitative outcomes (e.g., nap durations) were measured objectively by an accelerometer, whereas qualitative outcomes (e.g., difficulty going to bed or falling asleep) were subjectively measured by parent-report (40).

No two studies used the same outcome variables. All diet studies presented outcomes as categorical variables, with three studies investigating breastfeeding related outcomes (e.g., breastfeeding ≥4 months) (47, 48, 51), four studies investigating consumption of healthy and unhealthy foods (e.g. consuming sweetened beverages, salty snacks, or fruits/vegetables) (39, 45, 46, 50), and one study investigating between-meal eating (49). Two studies presented physical activity outcomes as categorical variables (e.g., high, medium, and low physical activity level versus sedentary) (39, 41), whereas one study used continuous variables (e.g., average accelerometer counts per minute). All sedentary behaviour variables were categorical (e.g., >4 versus ≤4 hours/day of TV viewing), and two out of three studies used TV viewing as a proxy for sedentary behaviour. There was a wide range of sleep variables, from number and duration of naps to variables relating to the quality of sleep.

Four studies investigated only one exposure and one outcome variable (41-43, 47). The remainder explored several outcome (40) or exposure variables (39, 44). Thus, the eight included studies reported on 63 associations between non-parental childcare and diet outcomes, nine associations with physical activity outcomes, three associations with sedentary behaviour outcomes, and 15 associations with sleep outcomes. Nine studies employed analytical strategies that accounted for potential confounding effects of other variables (i.e., multivariable regression models) (39, 41, 45-51), 8 of which investigated diet outcomes, whereas the remainder used simple statistical tests which did not account for potential confounding factors (40, 42-44). In the majority of cases, this was because the association between childcare and physical activity, sedentary behaviour or sleep was not the main focus on the paper.

### Synthesis of Findings

Table 2 presents detailed results for all relevant associations explored in each study.

**Table 2.**
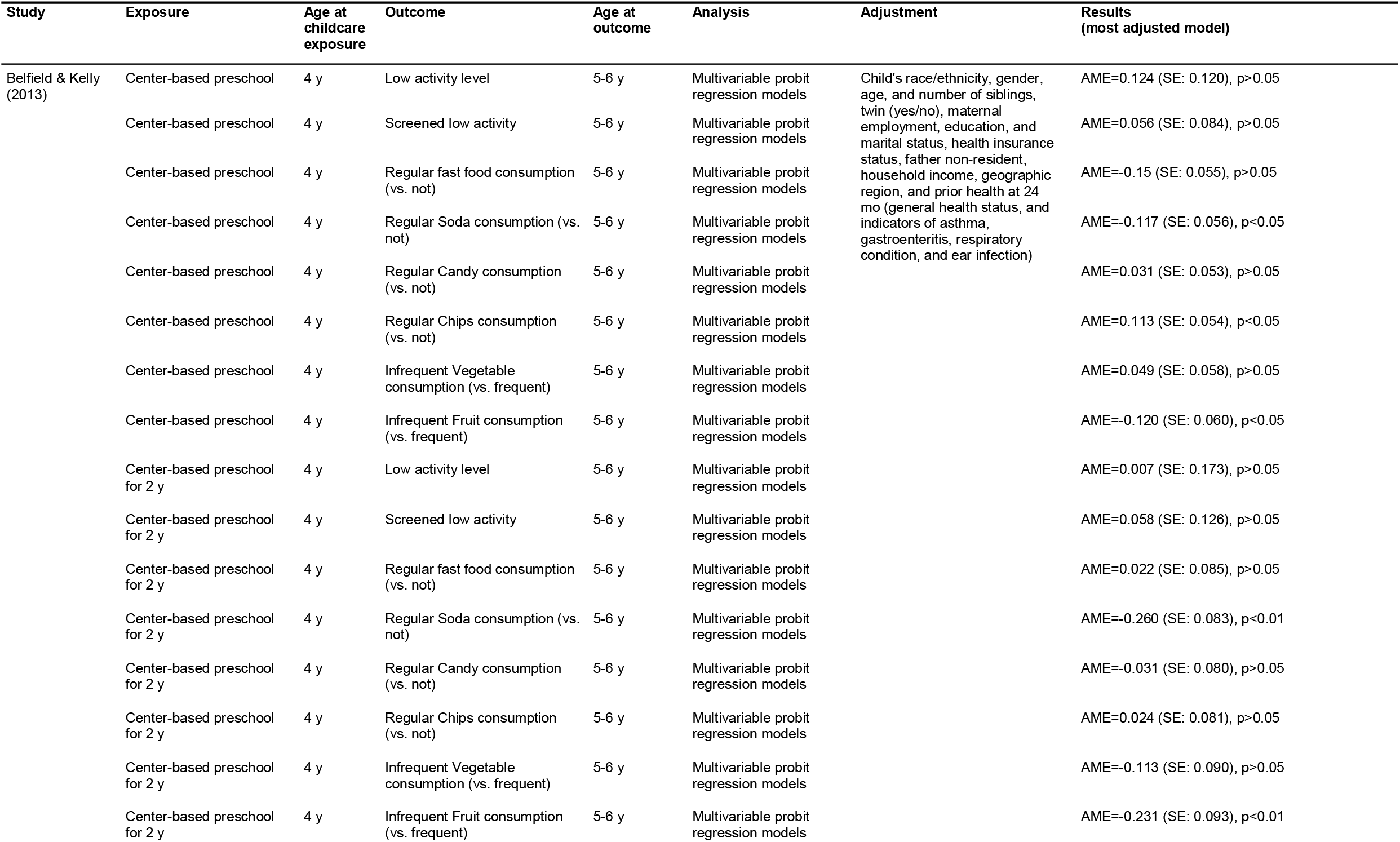
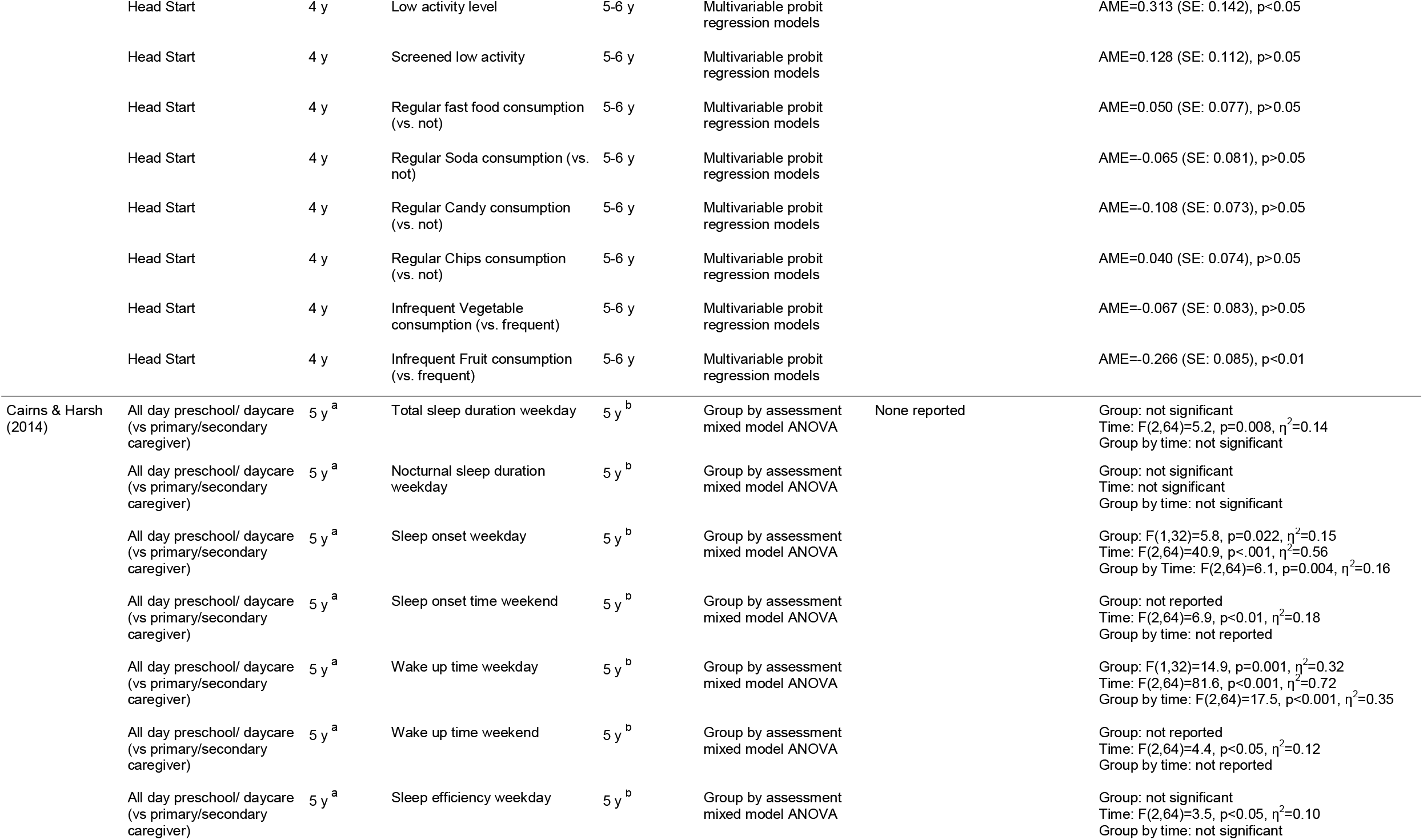
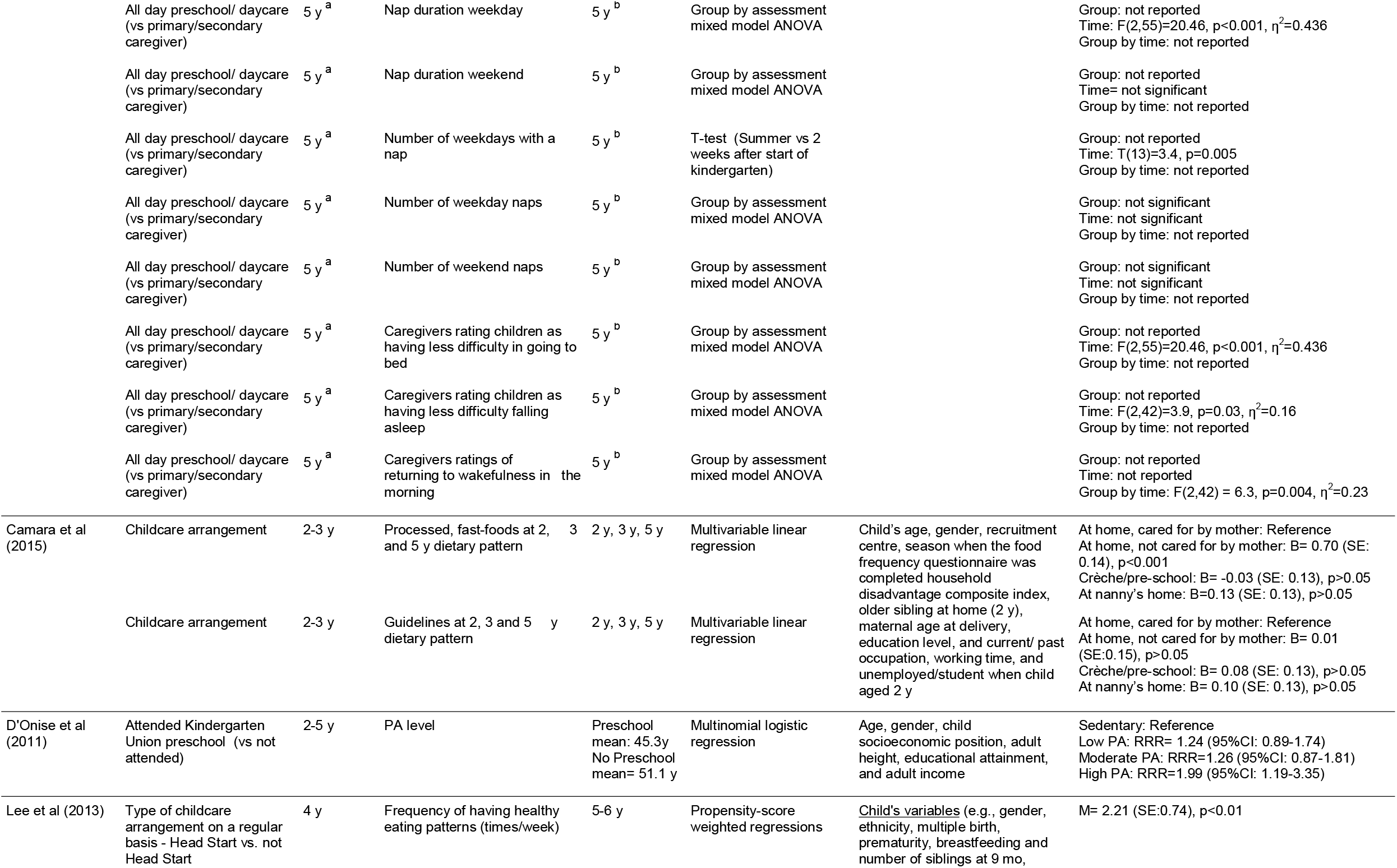
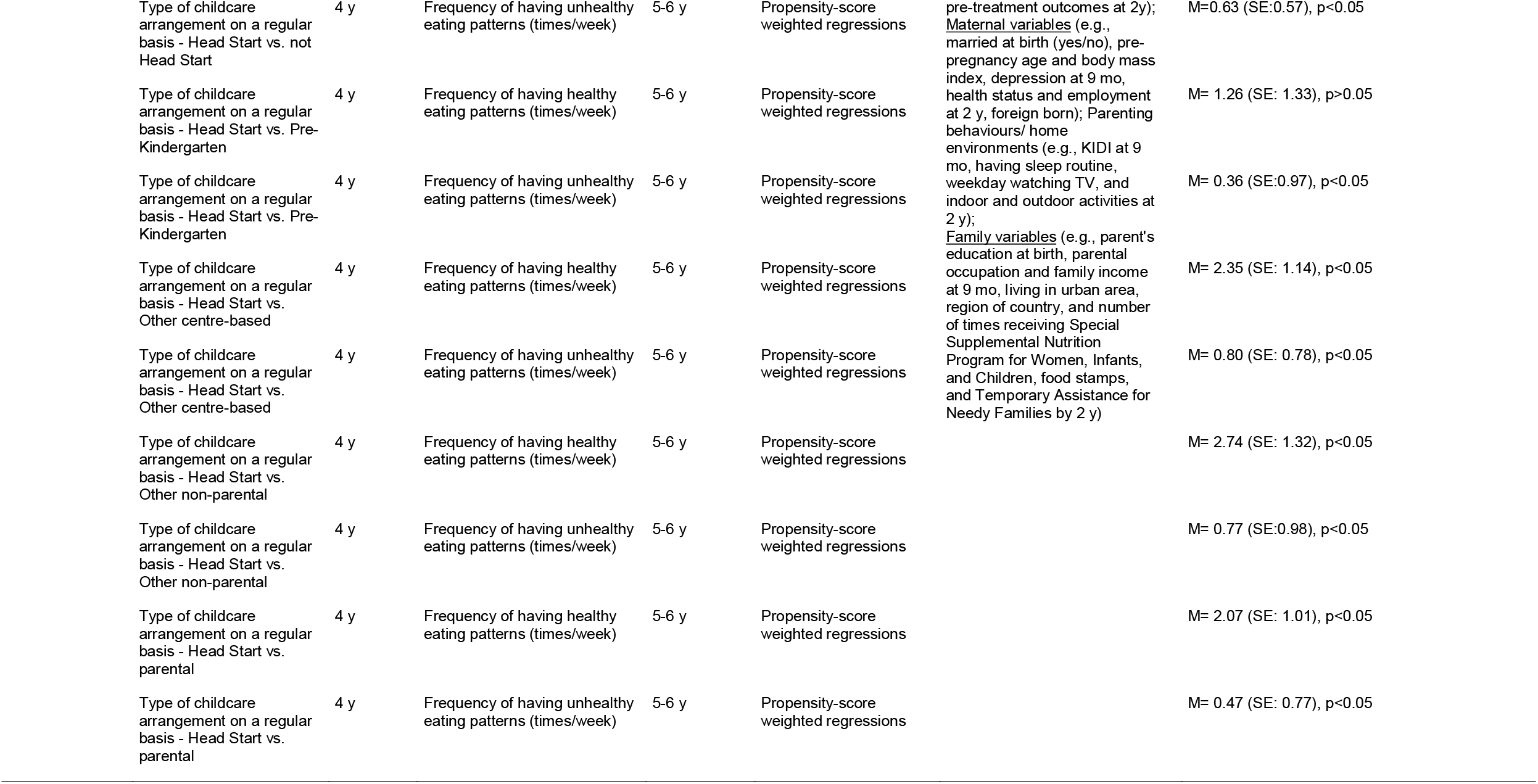
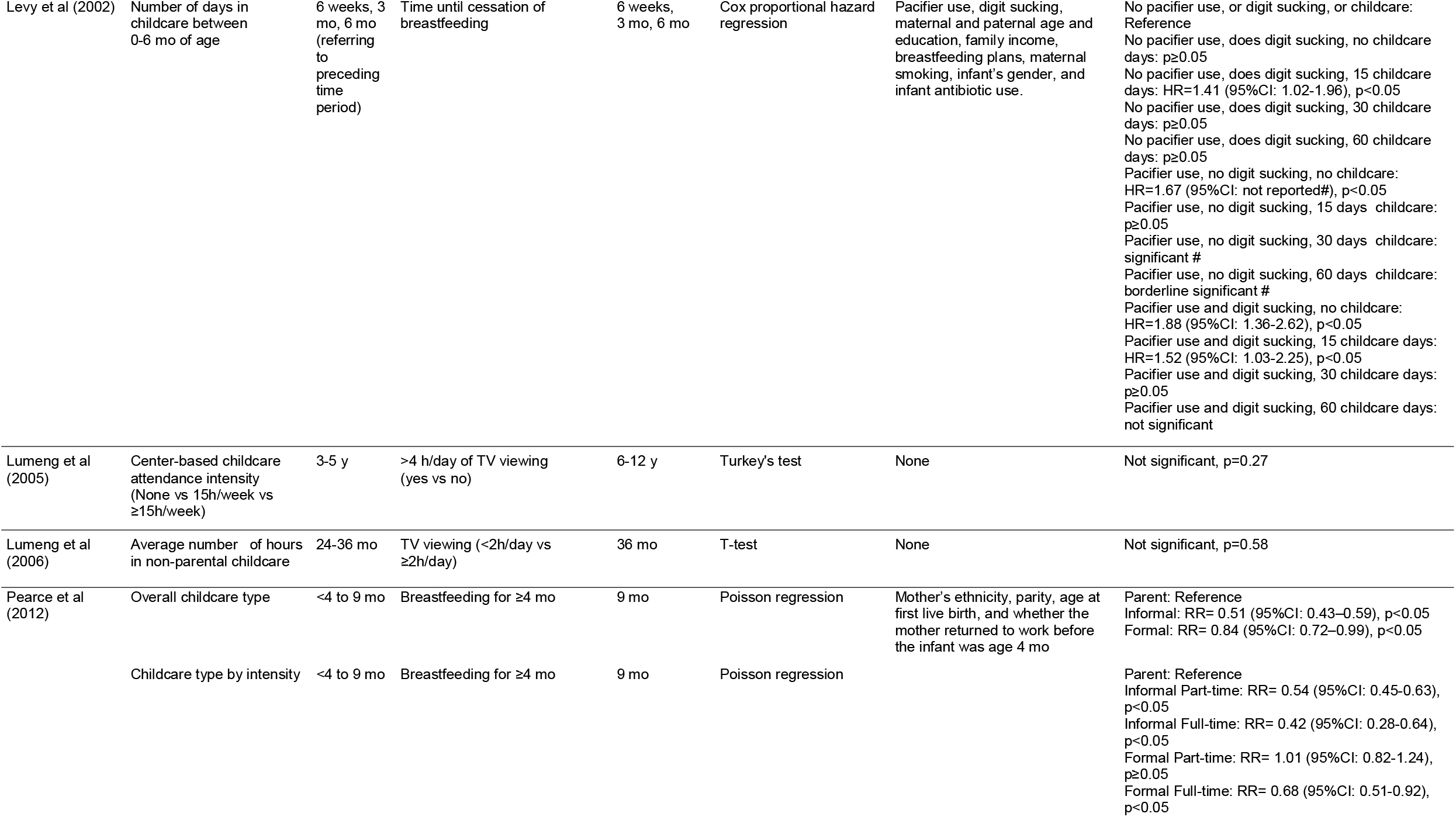
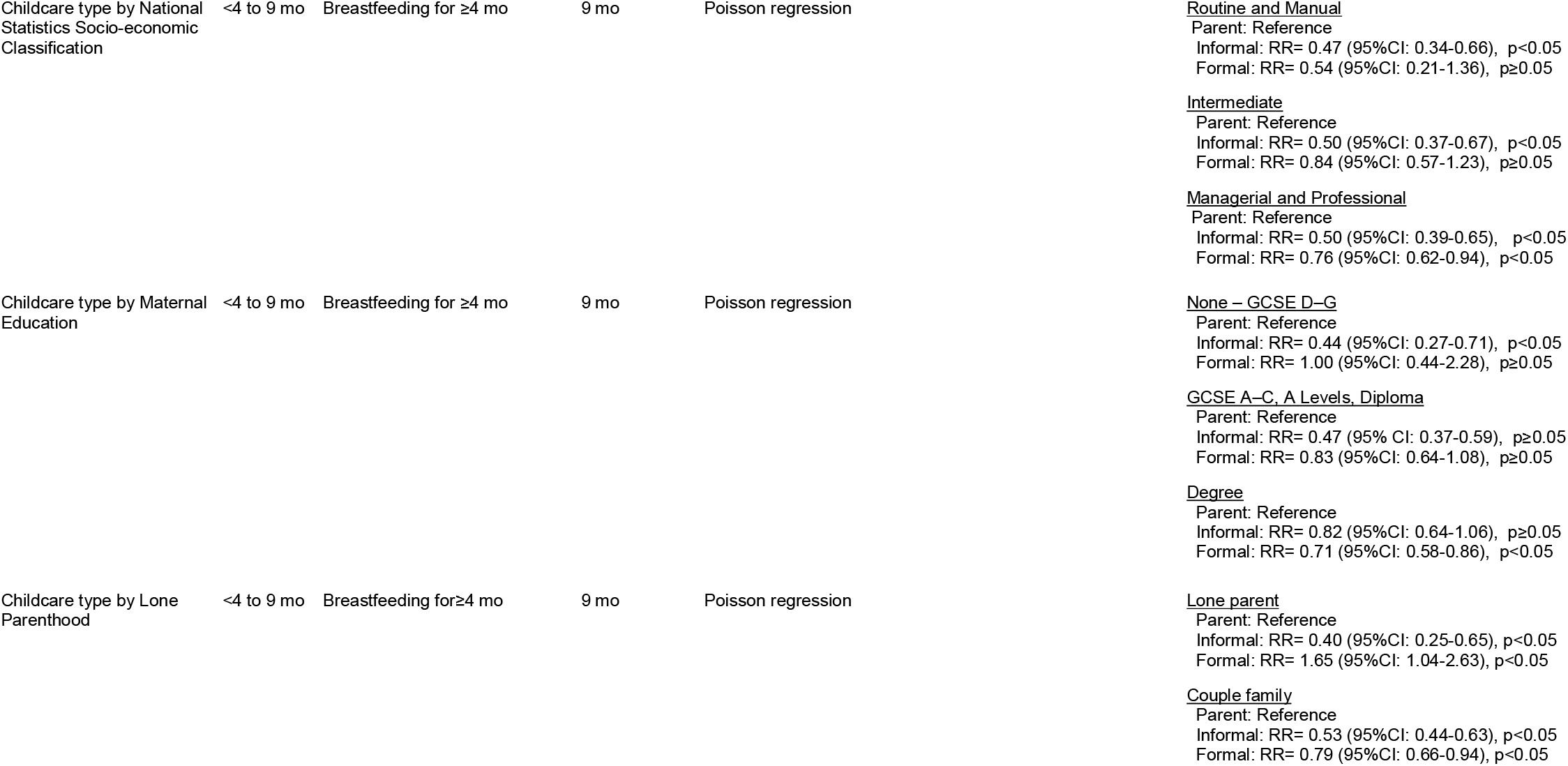
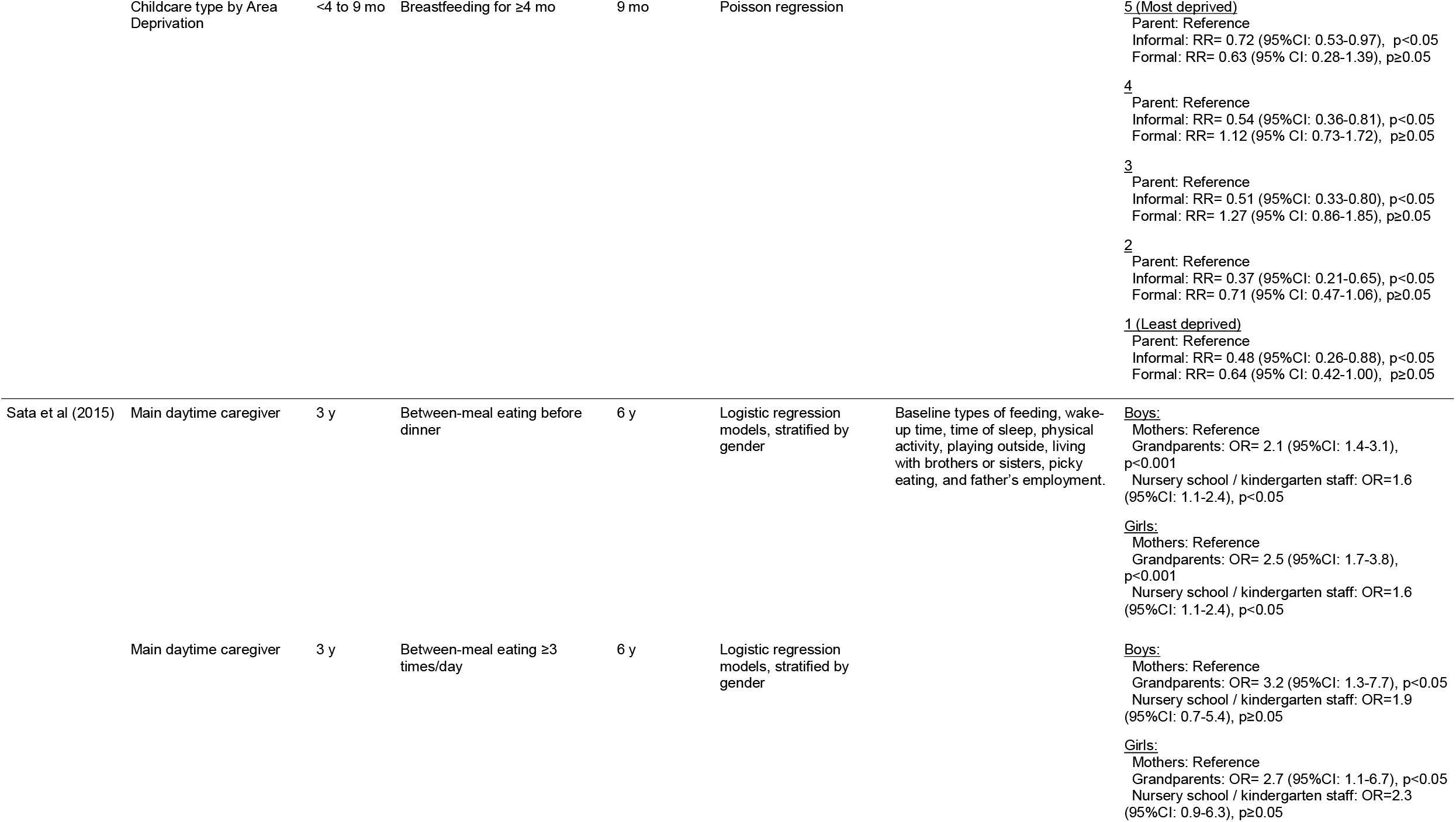
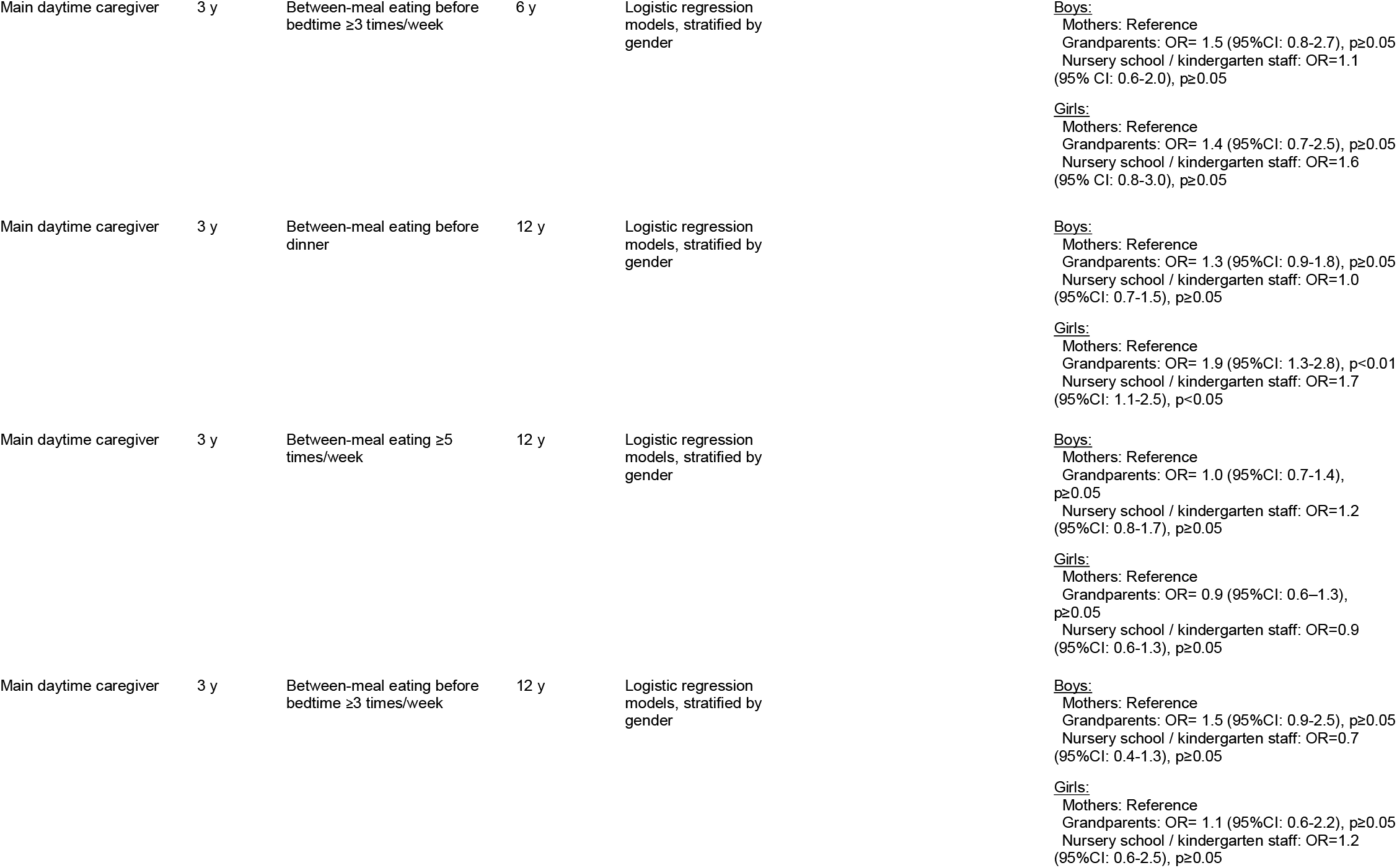
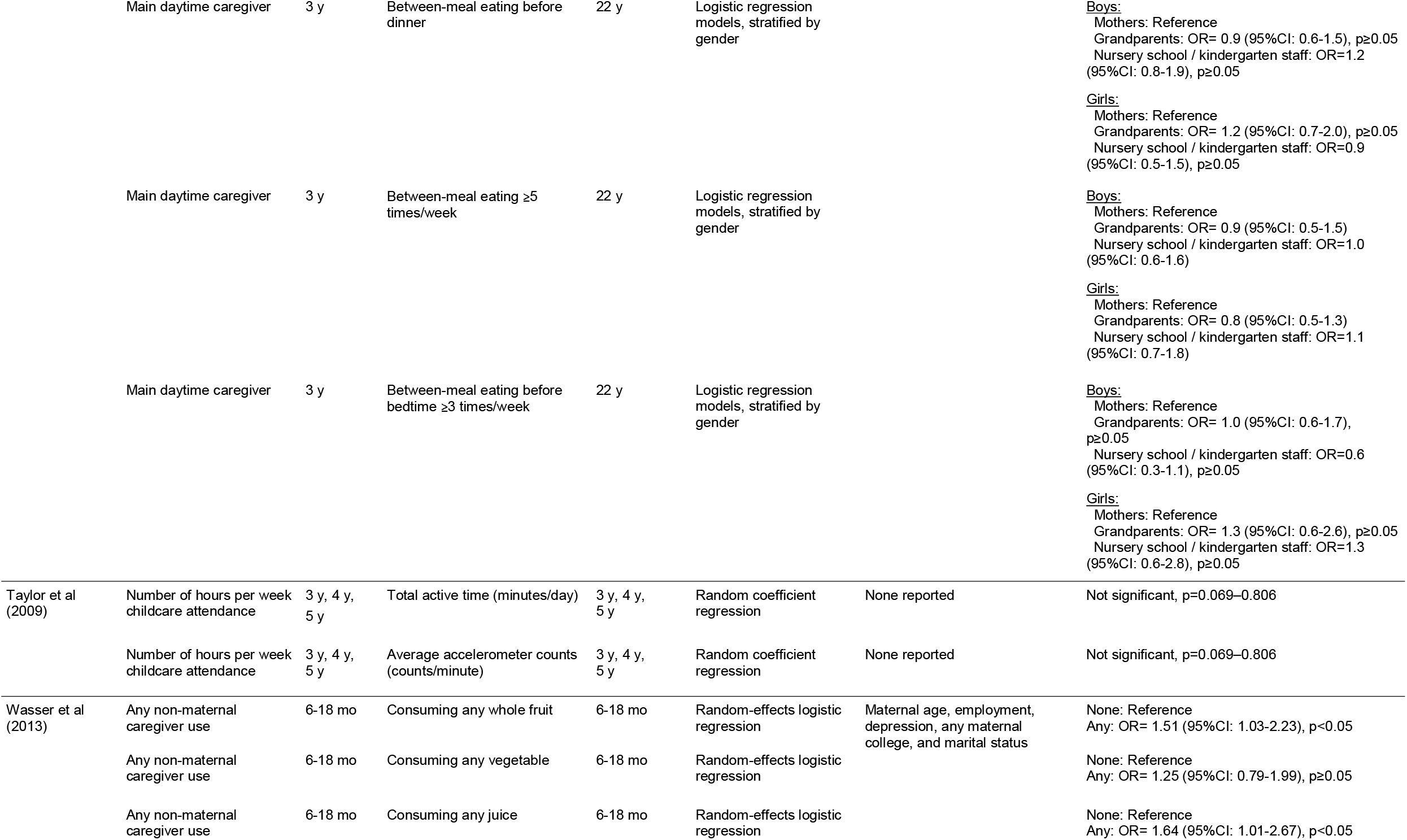
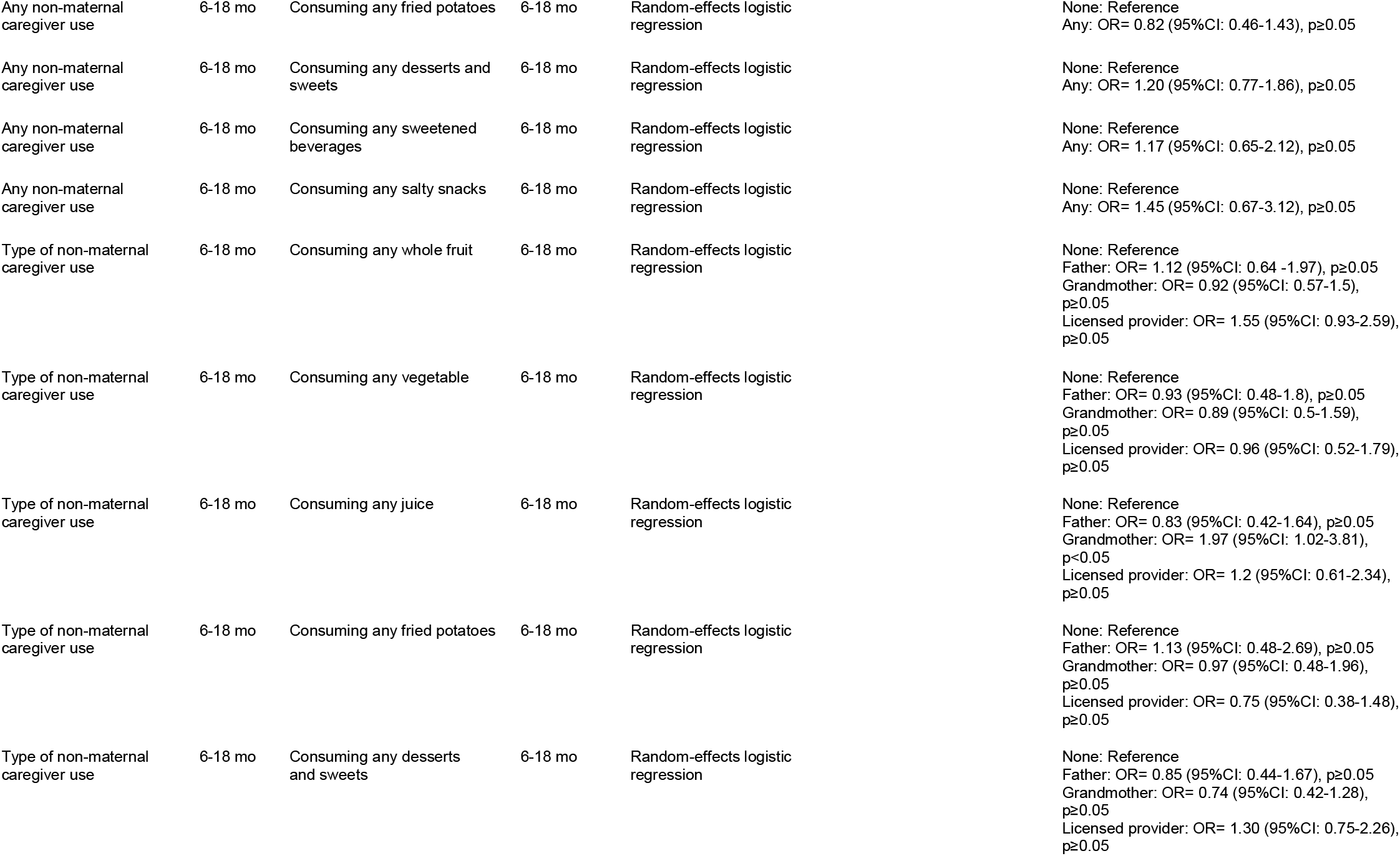
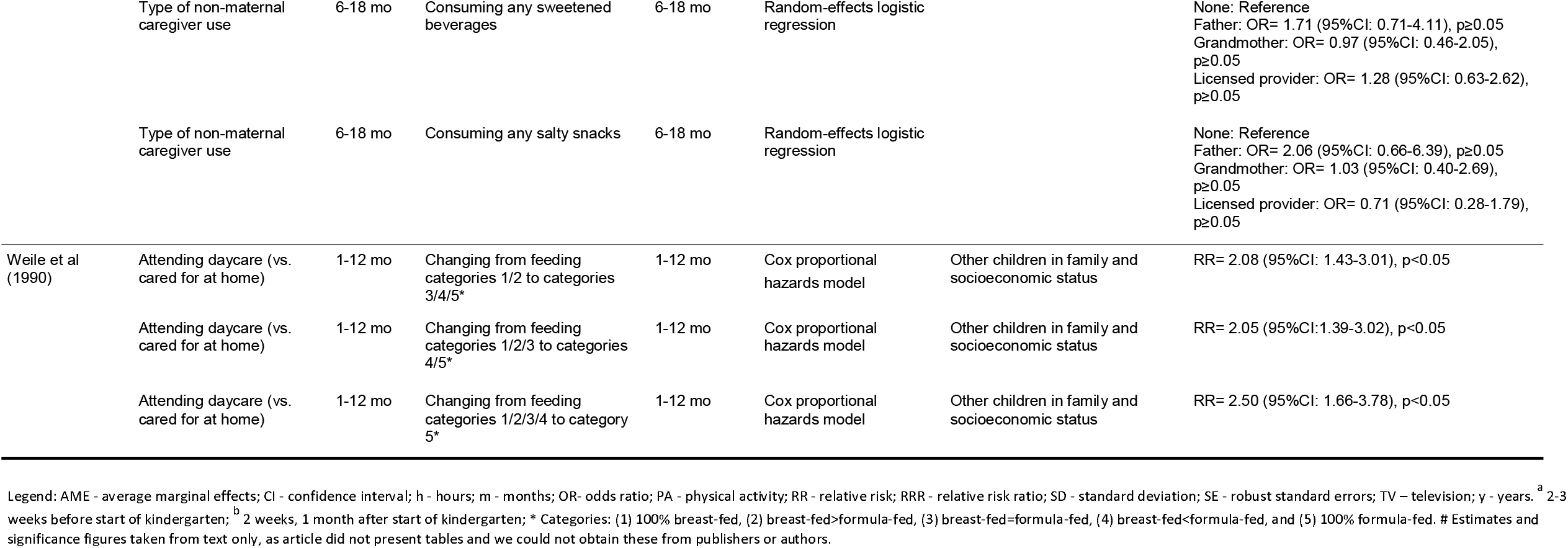
Results of included studies.

#### Diet

Eight studies evaluated the longitudinal relationship between non-parental childcare during early childhood and diet outcomes (39, 45-51). Results were highly mixed. Lee et al (46) reported that children who attended Head Start settings at 4 years of age showed significantly higher frequency of healthy eating patterns at 5-6 years of age than those attending other settings (all p<0.05), except Pre-Kindergarten. Conversely, no differences in frequency of unhealthy eating patterns were found between the groups (46). Another study assessing attendance at Head Start (39) found that children who attended Head Start or other centre-based childcare at 4 years of age (irrespective of length of exposure) were more likely to report frequent fruit consumption than those in other/mixed care (including parental care) at age 5-6 years. In this study, children who attended other (non-Head Start) centre-base childcare were also less likely to regularly consume soda at age 5-6 years than those in other/mixed care (all p<0.05). However, other centre-base childcare was also associated with higher likelihood of regular consumption of chips (p<0.05). There was no difference in the likelihood of regular consumption of fast-food, candy, and chips consumption, and frequent consumption of vegetables between those attending Head Start or other centre-based childcare (irrespective of length of exposure) versus other/mixed care (including parental care).

Similarly, Wasser et al (50) also reported mixed findings. They found that children in any non-maternal childcare had higher odds of consuming whole fruit (Odds Ratio (OR): 1.15, p<0.05), and juice (any childcare OR: 1.51; Grandparents OR: 1.91, p<0.05) than those in maternal care. But there was no association between childcare (overall or by type) and consumption of five other food and drinks including vegetables and salty snacks. Camara et al (45) also reported mostly null associations between childcare and the two dietary patterns investigated, except a higher adherence to a processed/fast-food pattern at 2-5 years of age in those being cared for at home by someone other than the mother compared to those cared for by their mother (B= 0.70 (SE:0.14), p<0.001) at 2-3 years of age. Sata et al’s (49) reported more frequent between meal eating before dinner at age 6 years in those cared for by grandparents and nursery/kindergarten than those cared for by mothers, as well as between meal eating ≥3 times per day those cared for by grandparents versus those cared for by mothers at 3 years of age. However, no other associations were found with any care at 12 or 22 years of age.

Three studies investigated breastfeeding outcomes (47, 48, 51), showing mixed results. Pearce et al (48) reported lower likelihood of breastfeeding for ≥4 months in children attending informal compared to parental care (independent of attending full-or part-time, lone parenthood, or area of deprivation), but mixed results for those attending formal care. For example, in the analyses stratified by family structure, children living in single parent families receiving formal care were more likely to be breastfed for ≥4 months (Risk ratio (RR)= 1.65) than those being cared for by parents, but the reverse was true for children living in couple families (RR= 0.79, all p<0.05). While Weille et al (51) reported a higher risk of changing from mostly breastfed to mostly or solely formula-fed in those attending childcare compared to those cared for at home (RR= 2.05 to 2.50, p<0.05), Levy et al (47) found an increased risk of earlier cessation of breastfeeding in children who used pacifier and did not attend any childcare and those who attended 15 days of childcare between 0-6 months of age versus those not attending childcare and not using pacifier.

Overall, the eight included studies tested 63 associations between non-parental childcare exposures and diet outcomes. Of these, 37 (59%) were null, 10 (16%) indicated significant beneficial effects of non-parental care on dietary behaviours, seven (11%) indicated significant detrimental effects of non-parental care on dietary behaviours, one (2%) found significant association with a diet behaviour that is of unsure detrimental effect (between-meal snacking) (49), and eight (13%) found mixed results, As an example of the latter, when investigating associations by maternal education, Pearce et al. (48) reported that those in informal care had significantly lower odds of being breastfed ≥4 months versus those in parental care, but only when parents had GCSE D-G or lower education. At the same time, those in formal care also had significantly lower odds of being breastfed ≥4 months versus those in parental care, but only when parents had degree-level education (48).

#### Physical Activity

Three studies evaluated the longitudinal relationship between childcare during early childhood and physical activity outcomes (39, 41, 44). Results were highly mixed. Belfield & Kelly (39) found that children who attended Head Start at 4 years old had significantly lower physical activity levels in kindergarten than those who received parental care. However, there was no difference in physical activity between those attending other centre-based care versus parental care, irrespective of length of exposure to such care. Conversely, D’Onise et al (41) reported that those attending Kindergarten Union preschool between 2 and 4 years old were more likely to be in the high physical activity level group (versus sedentary group) at around age 45 years than those who did not attend this preschool. In the only study looking at intensity of childcare use, Taylor et al (44) found no significant associations between weekly hours of childcare attendance at 3 or 4 years old and objectively measured physical activity 1 or 2 years later.

Overall, the six included studies tested nine associations between non-parental childcare exposures and physical activity outcomes. Of these, seven (78%) were null (39, 44), whereas two (22%) found significant differences but in competing directions, with one showing childcare to be associated with more, and one with less physical activity (39, 41).

#### Sedentary Behaviour

Three studies evaluated the longitudinal relationship between childcare during early childhood and sedentary behaviour outcomes (41-43), including one study that conceptualised sedentary behaviour as the absence of physical activity (also reported on above) (41). As noted, D’Onise et al (41) reported that those who attended Kindergarten Union preschool between ages 2 and 4 years were less likely to be in the sedentary group (versus the high physical activity level group) at around 45 years, than those who did not attend this preschool. The remaining two studies (42, 43) found no significant associations between number of hours per week of childcare at 24-36 months and 3-5 years and subsequent daily hour of television viewing at 36 months and 6-12 years respectively. Thus, three associations between non-parental childcare and sedentary behaviour were tested in included studies. Two (67%) were null (42, 43), and one (33%) showed a significant association between childcare and lower risk of sedentary behaviour (41).

#### Sleep

The only study investigating the longitudinal relationship between childcare and sleep outcomes yielded mixed results (40). Cairns and Harsh (40) reported that those attending all day preschool or day care at age 5 years (versus a primary/secondary caregiver) transitioned to earlier sleep onset and wake up time on week days in the first months of preschool. The authors are clear that the health implications of these differences are unknown. There were no differences between the groups in any other variables (e.g., difficulty in going to bed and nocturnal sleep duration on week days). Overall, 15 associations between non-parental childcare and sleep outcomes were tested in this single included study. The majority (n=13) were null, with only two showing significant results. The health implications of these are unknown.

### Quality Evaluation

Risk of bias scores ranged from 1-12 out of a total maximum of 26 (with lower scores indicating lower risk of bias) (see Table 3). The most common sources of bias were not reporting or using valid and reliable outcome measures (NEL-BAT question 13; 12 studies), and outcome assessors not blinded (or not clear whether they were blinded) to the intervention or exposure status of participants (NEL-BAT question 12; 10 studies). There was low risk of bias throughout in terms of inclusion and exclusion criteria (question 2), recruitment strategy (question 2), accounting for variations in the execution of the study from the proposed protocol or research plan (question 7), and length of follow-up across study groups (question 14).

**Table 3.** Results of the Nutrition Evidence Library Bias Assessment Tool risk of bias evaluation.

| NEL-BAT question | Belfield & Kelly (2013) | Cairns & Harsh (2014) | Camara et al (2015) | D'Onise et al (2011) | Lee et al (2013) | Levy et al (2002) | Lumeng et al (2005) | Lumeng et al (2006) | Pearce et al (2012) | Stata et al (2015) | Taylor et al (2009) | Wasser et al (2013) | Weile et al (1990) | Total score by question |
| --- | --- | --- | --- | --- | --- | --- | --- | --- | --- | --- | --- | --- | --- | --- |
| 1. Were the inclusion/exclusion criteria similar across study groups? | 0 | 0 | 0 | 0 | 0 | 0 | 0 | 0 | 0 | 0 | 0 | 0 | 0 | 0 |
| 2. Was the strategy for recruiting or allocating participants similar across study groups? | 0 | 0 | 0 | 0 | 0 | 0 | 0 | 0 | 0 | 0 | 0 | 0 | 0 | 0 |
| 5. Was there an attempt to balance the allocation between the study groups or match the study groups (e.g., through stratification, matching, propensity scores)? | 0 | 2 | 1 | 0 | 0 | 1 | 2 | 0 | 1 | 2 | 0 | 1 | 0 | 10 |
| 6. Was distribution of health status, demographics, and other critical confounding factors similar across study groups at baseline? If not, does the analysis control for baseline differences between groups? | 0 | 1 | 0 | 0 | 0 | 1 | 2 | 2 | 1 | 2 | 0 | 1 | 1 | 11 |
| 7. Did the investigators account for important variations in the execution of the study from the proposed protocol or research plan? | 0 | 0 | 0 | 0 | 0 | 0 | 1 | 0 | 0 | 0 | 1 | 0 | 0 | 2 |
| 8. Was adherence to the study protocol similar across study groups? | 0 | 0 | 1 | 0 | 1 | 1 | 0 | 0 | 1 | 1 | 0 | 1 | 1 | 7 |
| 9. Did the investigators account for the impact of unintended/unplanned concurrent interventions or exposures that were differentially experienced by study groups and might bias results? | 0 | 0 | 1 | 0 | 1 | 1 | 0 | 0 | 1 | 1 | 0 | 1 | 0 | 6 |
| 12. Were outcome assessors blinded to the intervention or exposure status of participants? | 0 | 1 | 1 | 1 | 2 | 2 | 0 | 0 | 1 | 2 | 1 | 1 | 2 | 14 |
| 13. Were valid and reliable measures used consistently across all study groups to assess inclusion/exclusion criteria, interventions/exposures, outcomes, participant health benefits and harms, and confounding? | 1 | 0 | 2 | 2 | 2 | 2 | 2 | 2 | 1 | 2 | 2 | 2 | 2 | 22 |
| 14. Was the length of follow-up similar across study groups? | 0 | 0 | 0 | 2 | 0 | 0 | 0 | 0 | 0 | 0 | 0 | 0 | 0 | 2 |
| 15. In cases of high or differential loss to follow-up, was the impact assessed (e.g., through sensitivity analysis or other adjustment method)? | 0 | 1 | 2 | 1 | 0 | 1 | 1 | 1 | 0 | 2 | 0 | 0 | 0 | 9 |
| 16. Were other sources of bias taken into account in the design and/or analysis of the study (e.g., through matching, stratification, interaction terms, multivariate analysis, or other statistical adjustment such as instrumental variables)? | 0 | 2 | 0 | 0 | 0 | 0 | 2 | 2 | 0 | 0 | 2 | 0 | 0 | 8 |
| 17. Were the statistical methods used to assess the primary outcomes adequate? | 0 | 0 | 0 | 0 | 0 | 0 | 2 | 2 | 0 | 0 | 2 | 0 | 0 | 6 |
| **Total study score:** | 1 | 7 | 8 | 6 | 6 | 9 | 12 | 9 | 6 | 12 | 8 | 7 | 6 |  |

## DISCUSSION

### Summary of findings

To our knowledge, this is the first systematic review to investigate the longitudinal relationship between non-parental childcare before age 6 years and diet, physical activity, sedentary behaviour and sleep. Overall, the evidence base is very limited with only 13 studies meeting eligibility criteria. In total, eight studies reported on diet outcomes, three on physical activity, three on sedentary behaviour and one on sleep. Included studies varied widely in terms of definition and measurement of both exposure and outcomes, and lacked in-depth exploration of different aspects of childcare that may influence any relationship with the outcomes studied. The available, limited, longitudinal literature suggests that attending certain types of non-parental childcare (particularly informal providers) might be related to less breastfeeding, but the evidence regarding other dietary outcomes is mixed, and sometimes contradictory. Moreover, the data reviewed suggests that attending non-parental childcare is unrelated to physical activity, sedentary behaviour or sleep outcomes. Included studies were of mixed quality with most (92%) not reporting using valid and reliable outcome measures, and few (23%) including blinding of outcome assessors to participants’ exposure status.

### Strengths and limitations of studies included in the review

The measurement of exposure to childcare in included studies was highly variable. Some studies focused on one particular type of childcare provider (e.g., attending Kindergarten Union) (41) versus a reference group that was an amalgam of all other types (39-41). Other studies included only the number of hours per week in non-parental childcare (42-44). Only four studies explored differences between the type of childcare provider (45, 48-50), but no studies performed detailed analyses exploring differences by duration, intensity, and timing of childcare. Thus, we were unable to explore differing effects according to these characteristics of childcare, as originally intended.

Similarly, there was substantial heterogeneity in how outcomes were assessed in included studies, which did not allow for direct comparisons. Apart from Cairns & Harsh’s study (40), no study reported validity or reliability of the methods used for measuring outcomes. Concurrently, the common use of proxy-report measures of diet, physical activity, sedentary and sleep behaviour outcomes increases the risk of measurement error and bias (e.g., overestimating physical activity and underestimating sedentary behaviour).

Only seven (54%) studies used an adequate analytical framework that accounted for the potential complexity of the relationship between non-parental childcare and outcomes (physical activity in both cases) (39, 41, 45, 48-50), by including and statistically adjusting for potential confounding and mediating variables. Thus, the evidence base may be substantially compromised by uncontrolled confounding by factors such as family and socioeconomic characteristics. Furthermore, apart from Pearce et al’s (48) study focusing on breastfeeding, no other study explored variations in any relationships between exposures and outcomes according to contextual factors, such as maternal education or socioeconomic status. Thus, we were not able to report on these, as originally proposed in the protocol (33). Failure to adjust for confounding variables is often a result of the association between childcare and health behaviours not being the primary focus of the study. Greater attention to these associations as primary aims of studies is required to increase the strength of available evidence.

### Strengths and limitations of the review

This systematic review has several strengths. A large number and variety of databases were searched using a comprehensive search strategy designed in partnership with an experienced librarian (VP), without limits on date of publication or language. Independent double screening was used at both abstract and full-text screening stages, and a third reviewer was included to resolve any inconsistencies in these processes, reducing risk of researcher bias. In cases of remaining uncertainty, study authors were contacted. The risk of bias assessment (NEL-BAT) was also performed independently in duplicate. This review focused on longitudinal studies because these provide a better indication of causality between exposure and outcomes than cross-sectional studies (52).

However, there are a number of limitations that need highlighting. The low number of studies for each of the outcomes did not allow us to present a summary of findings table as advised by the Cochrane Library Handbook, nor to perform a meta-analyses as planned (33). Heterogeneity in the study designs, definition of exposure and outcomes, and the methods and measurement tools used also made comparisons difficult; therefore, data were narratively synthesized and described. The results cannot be generalised to middle-income countries, as all studies were located in high-income countries. Furthermore, it was not possible to determine if associations persisted or emerged later, because only two articles and four (4%) associations had outcomes that were measured after childhood (41, 49); the remaining 11 articles and 85 (96%) associations assessed outcomes that were measured during childhood only (0-12 years).

### Interpretation of findings

Overall, there were substantial null results with a few scattered and mostly inconsistent statistically significant associations between non-parental childcare and diet, physical activity, sedentary behaviour and sleep outcomes. There was an indication that attending Head Start settings might be associated with positive dietary behaviours compared to other/mixed care (including parental care) (39, 46). However, this evidence comes from only from two studies (39, 46), and was not seen across all dietary outcomes studied (e.g., there was an association with more frequent fruit but not vegetable consumption) (39), or in relation to all other childcare types (e.g., no significant difference in healthy eating patterns between Head Start and state-funded pre-kindergarten programmes). Few consistent findings were found for physical activity, sedentary behaviour or sleep.

Whilst cross-sectional studies generally find more evidence of an unhealthy effect of childcare on diet and activity behaviours (22, 23, 26, 28, 30, 53, 54), this does not appear to be reflected in the limited available longitudinal data. It is possible that any cross-sectional relationship does not persist longitudinally and, hence, that there is no long-term impact of childcare on diet and activity behaviours. This would suggest that identified longitudinal associations between childcare and adiposity occurs via other mechanisms, such as stress. Alternatively, and maybe more likely, the quality and quantity of the longitudinal evidence available on the relationship between childcare and diet and activity behaviours is not strong enough to draw conclusions on the presence or nature of any relationship.

Most included studies measured outcomes in childhood, up to age 12 years, only (39, 40, 42-44). It is possible that any effects of childcare on diet and activity behaviours emerge later in life – particularly when children start to develop into more independent adolescents and adults. Although not enough to corroborate it, the significant associations found in D’Onise et al’s study (41), where physical activity level was assessed during mid-adulthood, support the plausibility of that hypothesis at least for activity behaviours.

The wide range of different outcome and exposure measures used in the included studies indicates poor theorisation and conceptualisation of any potential association. In general, there is limited evidence of shared understanding of exactly what aspect of childcare is expected to be associated with exactly what aspect of diet or type of activity behaviour, what the direction of such associations is, and why. Furthermore, authors rarely addressed the many dimensions that can vary in the exposure to non-parental childcare in terms of provider, timing, duration and intensity (particularly in relation to the activity behaviours outcomes). Greater conceptual clarity in these areas may help drive stronger longitudinal investigations. Clearer disentanglement of all of the potential dimensions in which exposure to childcare may vary will help identify if there are more and less healthful ways in which children can receive childcare.

Although it was not possible to perform meta-analyses or meta-regressions, there is no obvious indication that results were related to study size, whether outcomes were considered as continuous and categorical variables, whether outcomes were measured using subjective or objective methods, and whether studies were prospective or retrospective. However, the very small number of studies included for each outcome makes it difficult to draw conclusions on these issues.

### Implications for policy, practice and research

Although overall there was little evidence of a longitudinal relationship between childcare and diet and physical activity, sedentary behaviour and sleep, this more likely represents a current absence of high quality evidence, rather than good evidence of absence of an effect. Given this, it is difficult to draw any firm implications for policy and practice. Nevertheless, and given that there is some evidence of an association between childcare and adiposity,(15-17) it would be prudent for those regulating and providing childcare to continue to consider how they can provide a healthful environment for the children in their care.

The small number of studies included in the current review highlights the need for more longitudinal studies investigating the relationship between non-parental childcare and diet, physical activity, sedentary behaviour and sleep. These studies should employ valid and reliable measures of both exposure and outcomes; analytical frameworks that recognise the potential complexity of the relationship between exposure and outcome, and account for known and possible confounding and mediating factors (e.g., socioeconomic status and maternal employment). Additionally, studies should also perform more detailed investigations to explore potential differences in the effect of childcare according to the type of provider, duration, intensity and timing of childcare. This would help in clarifying whether specific patterns of exposure to non-parental childcare have a more or less healthful impact on children’s diet or activity behaviours.

The majority of studies included in this review assessed outcomes only during childhood (0-12 years), with only two studies assessing outcomes during adulthood (meal eating at age 22 years, and physical activity level at ages 45-51 years) (41, 49). There is a need for more studies examining long term relationships, to assess whether relationships between childcare and diet, physical activity, sedentary behaviour, or sleep emerge and continue into adolescence and adulthood. Existent birth cohorts may be useful in this respect, although they may impose limitations in the detail and methods of assessment of childcare exposure.

There is a lack of studies in middle-income countries, as well as consideration of differences in effect by ethnicity and socioeconomic status. Studies located in middle-income countries and investigating interactions with ethnicity and socioeconomic status would allow us to assess whether context influences the relationship between childcare and activity behaviours and hence whether targeted interventions may be justified. Indeed, the only included study which explored this aspect found that informal childcare was consistently associated with higher risk of not breastfeeding for at least 4 months across two indicators of socioeconomic status; but there were inconsistent, mostly null, associations with formal childcare (48).

## CONCLUSIONS

This review provides the first systematic summary of studies examining the longitudinal relationship between non-parental childcare and diet, physical activity, sedentary behaviour and sleep. Results were dominated by null findings with little consistent evidence that non-parental childcare was with any of the outcomes of interest. However, the available evidence is limited, highly heterogeneous in the definition and measurement of non-parental childcare, diet and activity behaviours, and lacks an in-depth exploration of different aspects of childcare that may influence this relationship, such as the type, duration or intensity. Further work is required to clearly conceptualise proposed pathways linking childcare with diet and activity behaviours, and to determine whether and what aspects of childcare might discourage physical activity, and promote sedentary behaviour, less healthful diet and sleep patterns. This would, in turn, help identify potential targets for interventions, policies, or regulations to help childcare settings provide healthful environments for the children in their care.

## Data Availability

Not applicable - systematic review

## DECLARATIONS

### Ethics approval and consent to participate

Not applicable.

### Consent for publication

Not applicable.

### Availability of data and material

Not applicable.

### Authors’ contributions

SC, SBN, and JA devised the systematic review, performed the risk of bias assessment, discussed and resolved disagreements on inclusion of studies at both title/abstract and full-text screening stages. VP advised on the design of the search strategy, conducted the database searches, and provided the search results for the review. SC lead on the merger and de-duplicating of the results database, and the drafting and submission of the manuscript. All authors performed screening at both stages, reviewed several drafts of the manuscript, and agreed on the final submitted version of the manuscript

## Acknowledgements

The authors thank Dr. Sarah Gonzalez-Nahm for her invaluable help in the double-screening process of this review, and Kai Schulze and Amy Yau for their help with translating and screening of non-English full-texts.

